# Effectiveness and acceptability of an opt-out nudge to promote influenza vaccination among medical residents in Nice, France: a randomized controlled trial

**DOI:** 10.1101/2022.09.09.22279772

**Authors:** Adriaan Barbaroux, Ilaria Serati

## Abstract

**Background:** Nudges have been proposed as an effective tool to promote influenza vaccination of healthcare workers. To be successful, nudges must match the needs of the target healthcare workers population and be acceptable.

**Objective:** To evaluate the effectiveness and the acceptability of an opt-out nudge promoting influenza vaccination among medical residents.

**Methods:** The hypothesis were that an opt-out nudge would be effective, better accepted when applied to patients than to residents, and that prior exposure to a nudge and being vaccinated increase its acceptability and residents’ sense of autonomy (the feeling of being in control of their choice about whether to get vaccinated). Residents were randomly divided into two parallel experimental arms: a nudge group and a control group. The nudge consisted in offering participants an appointment for a flu shot, while leaving them the choice to refuse or to reschedule it.

**Results:** The analysis included 260 residents. Residents in nudge group were more likely to be vaccinated than residents in control group. There was a strong consensus among the residents that it is very acceptable to nudge their peers and patients. Acceptability for residents and patients did not differ. Acceptability was better among residents exposed to the nudge and residents who were vaccinated. Residents considered that the nudge does not reduce their control over whether to get a flu shot. The sense of autonomy was associated with nudge’s acceptability.

**Conclusion:** An opt-out nudge to promote influenza vaccination among medical residents can be effective and very well accepted. These data suggest that this approach can complement other vaccination promoting interventions and be eventually extended to other healthcare workers’ categories and to general population, but should consider its ethical implications. More studies are needed to assess the nudge’s effectiveness and acceptability on other populations.

**Key messages:** Nudging is one of the most efficient techniques to improve vaccination coverage.

Changing defaults may be effective in promoting vaccination and well accepted.

Changing defaults does not lower the feeling of control over choices.

The sense of autonomy related to a nudge is correlated to its acceptability.

Behavior adoption increases the sense of autonomy related to a nudge.

Using defaults for nudge should take into account the ethical implications.

## Background

Influenza is a public health issue, with seasonal epidemics affecting 2 to 8 million people in France each year, with recent figures estimating up to 15.000 influenza-associated deaths(1). Healthcare workers are at risk of acquiring influenza and thus serve as an important reservoir for patients under their care. Also, they are in contact with those who are most at risk of severe influenza(2). Annual influenza vaccination of high-risk persons and their contacts, including healthcare workers, is a primary means of preventing influenza, limiting the use of care and reducing pneumonia deaths in health facilities(2).

Despite influenza vaccine effectiveness and safety, vaccination coverage among healthcare workers in Europe rarely exceeds 30%(3–5) and remains far below recommended levels of 75%(6).

Most public health policies use education programs to change healthcare workers’ attitude and increase influenza vaccine uptake(7). The effects of conventional educational programs and campaigns are in general of modest impact only, suggesting that new strategies to promote influenza vaccination among healthcare workers are needed(8).

Thaler and Sunstein suggested that nudges are an effective method to promote behavior changes(9). A nudge refers to any alteration of the environmental context or “choice architecture” people operate within, that aims to influence people’s behavior in a predictable way, without denying them any options or changing their attitudes(9). It consists of a gentle incitement that respects freedom of choice and does not use financial incentives(9). Its effectiveness has been demonstrated in the field of economics and earned Richard Thaler the Nobel Prize in Economics in 2017.

Nudges’ effectiveness in medicine is poorly studied, but systematic reviews place nudges among the most promising interventions to promote healthy behaviors(10,11). Some studies have shown nudges’ effectiveness in increasing vaccination coverage(11–19).

There are different types of nudges, which change the choice architecture in different ways(20). Patel et al.(18) showed that the most effective nudge consists in changing the default (the outcome that results when no action is taken) by scheduling people for a vaccination appointment, making the appointment the default option (opting-out), and thus avoiding the risk of forgetting or procrastinating to schedule an appointment by oneself (opting-in).

According to Thaler and Sunstein, nudges must be transparent and publicly defensible(9). Being often very subtle, nudges might be criticized as unethical(21). Also, the operationalization of nudges is based on heuristics and cognitive bias(22). The analysis of the decision process induced by nudges shows that their handling is ethically tricky(23). By not being transparent about the intention to influence individual choice, they might be perceived as limiting freedom of autonomous decisions(21,24) or as manipulation attempts(25,26). Previous studies described a generally positive but variable acceptability, with 40 to 87% of participants judging nudges as acceptable(27). A nudge’s acceptability could also be modified by mere exposure to nudge(28).

Although there are several types of nudges, few studies evaluated their respective effectiveness, and it is difficult to predict a nudge’s effectiveness(29). Systematic reviews emphasize the need to develop better quality randomized trials(30). Also, there are few randomized controlled studies evaluating the acceptability of nudges(12,31,32) or their implementation among health professionals(11,30,33).

In a previous study on a medical residents’ population, a reminder nudge proved ineffective but well accepted(31). The aim of the present study was to evaluate the effectiveness and the acceptability of an opt-out nudge to promote influenza vaccination among a very similar residents’ population. It was hypothesized that the vaccination rate would be higher among nudge-exposed residents than in unexposed residents (nudge’s effectiveness, H1). The hypothesis about nudge’s acceptability were: acceptability would be higher when the nudge is applied to patients than to peers (H2), among nudged participants (H3) and those who were vaccinated (H4); the sense of autonomy (participants’ feeling of control over whether to get vaccinated) would be greater among nudged participants (H5) and those who were vaccinated (H6); the sense of autonomy would be positively correlated to the nudge’s acceptability (H7).

## Methods

### Setting, participants, design and procedure

This paper is an extension of a previous work testing the same hypotheses on a different nudge(31). Pre-recorded hypotheses are available on ClinicalTrials.com (ClinicalTrials.gov Identifier: NCT03768596).

A monocentric, controlled, randomized, two arm trial was designed. The inclusion criteria were to be a medical resident at the University Côte d’Azur and to be in an internship in one of the followings: Antibes Hospital, Grasse Hospital, Fréjus Hospital, the University and Teaching Hospital of Nice (Archet, Pasteur or Cimiez Hospital), or an ambulatory internship in Nice or at most 10 km from Nice. These internship sites rely on vaccination centers whose operators approved the implementation of the study. The exclusion criteria were the refuse to participate, the impossibility to reach the resident by phone, and accidental cross-over between the groups. An equal randomization (1:1) was used, stratified by internship site to avoid the center-effect due to discrepancies in vaccination campaigns in different hospitals.

The study was conducted in two steps. Step 1 took place between November 14^th^ and 27^th^, 2021. In step 1, residents assigned to the nudge group received a phone call from the main investigator (IS) to offer them an appointment for an influenza vaccination at the vaccination center on which they depended according to their internship site, while leaving them the choice to refuse or to reschedule it. No reference to the study was made at this stage. The control group was not solicited at that time and was included directly in step 2. Step 2 took place between January 9^th^ and February 28^th^, 2022. In step 2, both groups received a phone call from the investigator. The call specified its scientific purpose and the legal context for collecting data (anonymous, computerized processing, no obligation to reply, possibility of leaving the study at any time). The participants who gave a verbal consent answered a questionnaire about their vaccination status, their opinions on vaccination and their attitude towards the nudge (social acceptability and feeling of control). The questionnaire included an explanation of the nudge procedure and unveiled the study aim.

Participants self-reported their vaccination status by indicating whether they had obtained an influenza vaccination by January 9^th^, corresponding to the beginning of step 2. Social acceptability was assessed for the residents and for the patients via a series of seven-point Likert scales already used in a previous work(31), formulated as follows: “What do you think about the use of this type of method on you?” on a scale from 1 (not at all) to 7 (absolutely) and applied to the following eight adjectives: abusive, acceptable, inappropriate, ethical, immoral, unfair, legitimate, relevant. Acceptability toward patients was assessed via the same scales. Sense of autonomy was assessed via a seven-point Likert scale formulated as follows: “Do you think this nudge lets you choose whether to get vaccinated?” on a scale from 1 (not at all) to 7 (absolutely).

The telephone interview guide for step 1 and step 2 calls and the questionnaire are available online (Annex 1 and 2).

In order to obtain a statistical power of 90% with a tolerated alpha risk of 5%, 108 residents per group were required for the effectiveness study, assuming 90% vaccinated residents in the nudge group against 73% in the control group, the latter corresponding to the influenza vaccination rate found in our previous study(31). To obtain the same power for the acceptability study, 33 people were needed per group by providing an acceptability of 5 points on a Likert scale of 7 in the nudge group versus 4 in the control group.

### Data analysis

The effectiveness of the nudge on vaccination rates were evaluated by Chi2 tests. The consistency of the nudge acceptability scales was tested using Cronbach’s alpha.

The acceptability of the nudge for residents and for patients was compared using Student’s t.

The hypotheses on nudge’s acceptability and sense of autonomy were tested by stepwise linear regressions including the following independent variables: nudge exposure, age, gender, vaccination status, and opinion of current vaccination recommendations.

The dependent variables were vaccination status in step 2 (for the nudge’s effectiveness assessment), nudge’s acceptability for residents and patients, and sense of autonomy.

All statistical analyses were performed using JASP® software(34).

### Regulatory and ethical aspects

The study was classified out of scope of the Jardé law by the Ethics Committee “CPP Sud Ouest et Outre Mer III” (ID-RCB: 2021-A00975-36, Ref. SI CNRIPH: ID12087

N°21.03.31.68613). A favorable opinion was issued by the Ethics Committee of the French National College of Generalist Teachers (N°020921306). This work has been the subject of a declaration of compliance with the MR004 to the National Commission of Liberty and Computing “CNIL” (N°2221800v0).

## Results

### Descriptive statistics

Step 1 (inclusion and nudge application) took place between November 14^th^ and 27^th^, 2021 and step 2 (data collection) between January 9^th^ and February 28^th^, 2022. Of the 329 eligible residents, 46 could not be reached, 22 refused to participate and 1 was excluded because of accidental cross-over (missing data: 21%). Flow chart is available in Figure 1. The final analysis included 260 participants distributed as follows: 138 participants in the nudged group and 122 in the control group. Participants’ age ranged from 21 to 43 years (*M* = 26.8, *SD* = 2.8); 140 were women (67.5%).

**Figure 1.**
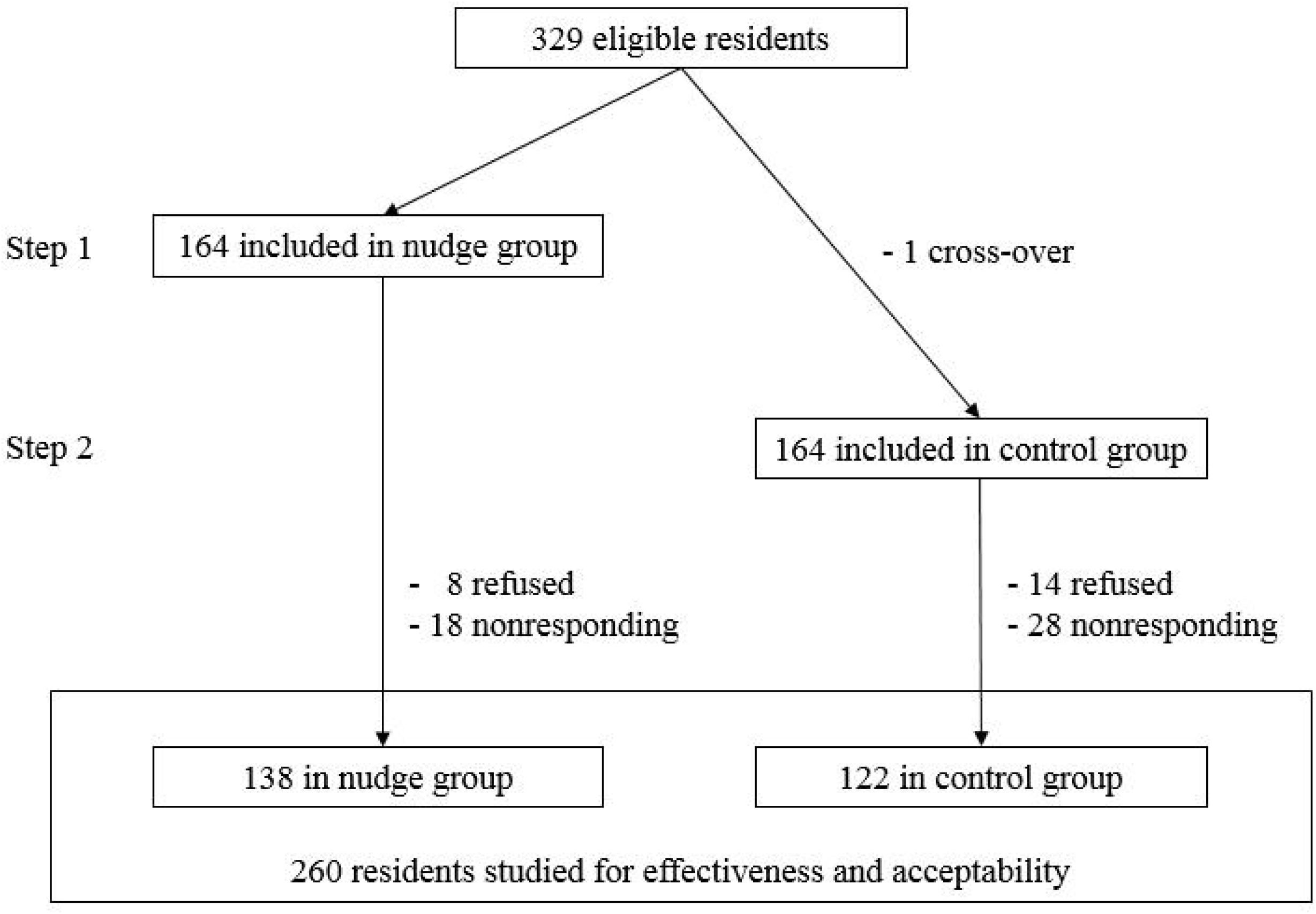
Flow chart.

**Figure 1a.**
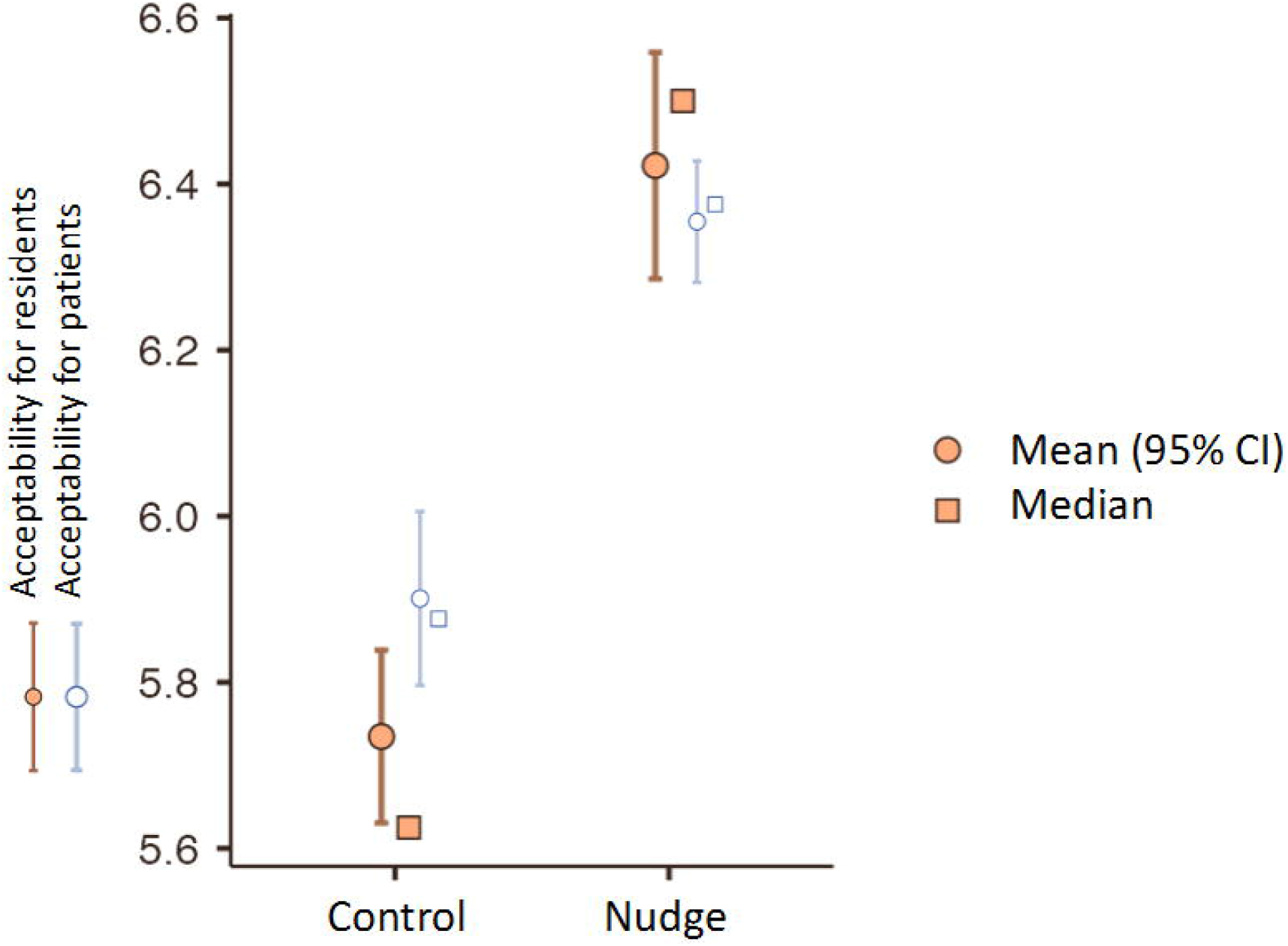
Mean acceptability of a nudge as applied to patients or to residents, among 260 residents from Nice, France in 2022 (error bars displays 95% CI).

Of the 164 participants included in step 1, 11 (6.7%) were already vaccinated at the time of inclusion. In step 2, 220 participants reported being vaccinated (84.6%). Table 1 presents the number of vaccinated participants per group and percentages among 260 residents.

**Table 1.**
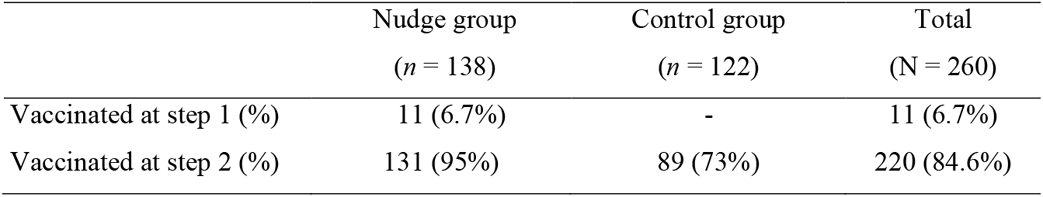
Number of vaccinated participants per group and percentages for residents from Nice, France in 2022 (N = 260).

### Nudge effectiveness

The first hypothesis (H1: residents exposed to the nudge were more likely to be vaccinated than unexposed residents) was verified, *χ*^2^(1, *N* = 260) = 70, *p* < .001.

### Nudge acceptability

We observed a high level of acceptability regardless of the experimental condition the residents were included in (see Table 2). Participants rated the nudging procedure acceptable for both patients (*M* = 6.14; *SD* = .56) and residents (*M* = 6.10; *SD* = .80). Only one resident considered the nudge procedure to be rather unacceptable when applied to residents or patients (acceptability below the seven-point Likert-scale midpoint).

**Table 2.** Acceptability of the nudge when applied to residents or patients per group and per vaccination status for residents from Nice, France in 2022 (N = 260).

| Acceptability | Nudge group | Control group | Total |
| --- | --- | --- | --- |
| For residents |  |  |  |
| Vaccinated | 6.46 (0.82) | 5.87 (0.54) | 6.22 (0.78)<sup>†</sup> |
| Not vaccinated | 5.79 (0.23) | 5.38 (0.56) | 5.45 (0.54)<sup>†</sup> |
| Total | 6.42 (0.82)<sup>*</sup> | 5.73 (0.59)<sup>*</sup> | 6.10 (0.80) |
| For patients |  |  |  |
| Vaccinated | 6.38 (0.44) | 6.03 (0.53) | 6.24 (0.50)<sup>††</sup> |
| Not vaccinated | 5.95 (0.22) | 5.55 (0.63) | 5.62 (0.59)<sup>††</sup> |
| Total | 6.47 (0.61)<sup>**</sup> | 5.90 (0.59)<sup>**</sup> | 6.14 (0.56) |

Values in brackets correspond to the SD. Results marked with \*, \*\*, † and †† were significant ( $p < .001$ ) and correspond to our pre-registered hypothesis.

The second hypothesis (H2: better acceptability for patients than for residents) was not verified: acceptability for residents and patients did not differ, *t*(260) = -1.18, *p* = .24.

The third (H3) and the fourth (H4) hypothesis were verified: residents were more accepting of the nudge as applied to themselves or to patients after previous exposure to the same nudge (*p* < .001) and when they were vaccinated (*p* < .001). See Figure 2 for a visual representation of these data and Table 2.

### Sense of autonomy

We observed a high level of sense of autonomy regardless of the experimental condition the residents were included in (see Table 3). No resident considered the nudge procedure to lower their control (sense of autonomy below the seven-point Likert-scale midpoint). The fifth (H5) and the sixth (H6) hypothesis were verified: residents felt more control over the nudge after previous exposure to the same nudge (*p* < .001) and when they were vaccinated (*p* < .001). The seventh hypothesis (H7) was verified: the sense of autonomy was associated with nudge acceptability as applied to the residents (*p* < .001) and patients (*p* < .001).

**Table 3.** Sense of autonomy related to an opt-out nudge for residents from Nice, France in 2022 (N = 260).

|  | Nudge Group<br>(n = 138) | Control Group<br>(n = 122) | Total<br>(n = 260) |
| --- | --- | --- | --- |
| Vaccinated | 6.81 (0.41) | 5.83 (0.93) | 6.41 (0.83)<sup>†</sup> |
| Not vaccinated | 6.86 (0.38) | 4.88 (0.78) | 5.22 (1.05)<sup>†</sup> |
| Total | 6.81 (0.41)<sup>*</sup> | 5.57 (0.99)<sup>*</sup> | 6.23 (0.96) |

Values in brackets correspond to the SD. Results marked with <sup>\*</sup> and <sup>†</sup> were significant ( $p < .001$ ) and correspond to our pre-registered hypothesis.

### Additional analyses

Residents were more accepting of the nudge when they approved vaccination recommendations or when they would recommend vaccination to patients or other healthcare workers. Acceptability did not differ within age and sex of the participants.

## Discussion

These data show that an opt-out nudge can increase medical residents’ influenza vaccination coverage. Also, it was perceived as a soft and acceptable incentive to target both residents and patients and did not reduce participants’ sense of autonomy (the feeling of control over whether to get vaccinated). Prior exposure to the nudge increased its acceptability and participants’ sense of autonomy. Acceptability and sense of autonomy were stronger among vaccinated participants.

### Effectiveness

In this randomized, controlled experimental study, a nudge changing the default (opting-out instead of opting-in for a flu shot) increased influenza vaccination coverage among medical residents. This result is consistent with previous evidences, which place opt-out nudges among the most effective interventions in promoting vaccination(14–18).

This result is even more remarkable when considering the high vaccination coverage of the study population, which is close to the recommended levels(6). Previous works showed that vaccination coverage is typically lower among medical residents(4).

The vaccination booths set up at the hospitals act like a nudge changing the option-related efforts, by facilitating the access to vaccination. The present study shows that this opt-out nudge can increase the expected benefits of other interventions aimed at increasing influenza vaccination coverage of medical residents.

### Acceptability

This study showed interesting results concerning the acceptability of this kind of nudge. First, the nudge was widely accepted, to a greater extent than previous works suggested(27,35). Second, there was a strong consensus among the residents that it is very acceptable to nudge both their peers and patients. Third, nudge exposure had a positive impact on nudge’s acceptability, as previously shown(31). Fourth, vaccinated residents found the nudge more acceptable than unvaccinated residents, according to recent evidences(32).

Both exposed and unexposed residents found the nudge very acceptable, with almost all residents accepting the nudge despite the embedded deception in the experimental design. Indeed, only at a later stage were participants informed of the experimental intent of the proposed vaccination appointment, which was actually a nudge aimed at influencing their vaccination behavior. This shift in the communication contract could have been perceived as a manipulation attempt(25), but it was not interpreted this way by participants. These findings suggest a wide acceptance supporting vaccination and interventions to promote it and are consistent with recent evidences about acceptability of nudges to promote vaccination on residents and healthcare workers(31,32). On the contrary, this massive acceptability was not found in general, non-medical populations(27,35). This discrepancy confirms that the population and behavior studied affect nudge’s acceptability, as previously evocated(31). Indeed, despite vaccine hesitancy among healthcare workers(36), most agree that vaccination is one of the most important public health practices(37). Residents and healthcare workers may be more inclined to accept a nudge about vaccination than general, non-medical population.

### Sense of autonomy

Despite nudge’s effectiveness, residents did not report any attempt to their sense of autonomy. Participants’ sense of autonomy was stronger in nudge-exposed and in vaccinated residents, and it was associated with nudge’s acceptability. Interestingly, residents for whom the nudge was effective felt they had more control over their choice than the others.

Participants were offered a vaccination appointment without explaining them it was a nudge. This lack of transparency could be experienced as a violation of freedom of choice(21,24), but it was not interpreted that way. This inconsistency can be explained by study’s conditions. Indeed, the design of the nudge made it very easy to opt out. This was found to be a relevant condition that needs to be considered when using defaults to influence health-related behaviors(38). To our knowledge, no experimental study has focused before on assessing people’s sense of autonomy related to a nudge.

### Strengths and limitations

One limitation concerns the method by which efficacy was assessed. To avoid measurement bias related to the expected long duration of the data collection phase (51 days), it was decided to collect the participants’ vaccination status on January 9^th^, 2022, which corresponds to the beginning of step 2. The influenza vaccination campaign lasted until February 28^th^, 2022(39). We assume that this may have underestimated vaccination coverage. However, the difference between the vaccination coverage of the two groups is so marked (22%) that we believe it is unlikely that such a difference can be due only to late vaccinations, taking into account that the vaccination campaigns began between late October and mid-November 2021 in all the hospitals involved.

In addition, since vaccination status was defined on declarative data and most hospitals did not kept track of employees’ influenza vaccinations, it was not possible to verify statements about vaccination status. Except for those who were already vaccinated at step 1, none of the participants in the nudge group refused the vaccination appointment. Since they were announced that they were free to accept or not, we have no reason to think that participants in the nudge group might have given more misinformation about their vaccination status than participants in the control group. While an overestimation of the vaccination coverages is possible, we do not believe that this could affected the assessment of the nudge’s effectiveness.

The strength of this study resides in its interventional, controlled and randomized design. Indeed, systematic reviews showed that, despite the numerous publications on nudges, only a few authors used the consort reporting guidelines as we did(29,30,33). Moreover, experimental studies conducted on the effectiveness and acceptability of a nudge are scarce(31). To our knowledge, this is the first attempt at studying participants’ participants’ sense of autonomy related to a nudge in experimental conditions, showing that the sense of autonomy is associated with nudge’s acceptability and that prior exposure to a nudge influence people’s sense of autonomy.

## Conclusion

This work shows that a nudge changing the default from opting-in to opting-out for a flu shot appointment is effective to increase influenza vaccination coverage among medical residents. Also, it was deemed a very acceptable intervention to promote vaccination of both residents and patients. This procedure did not reduce people’s feeling of control over their choices. Assessment of sense of autonomy is innovative in this context and deserves further investigations. Indeed, peoples’ sense of autonomy is correlated with nudge’s acceptability.

These data suggest that changing the default to promote influenza vaccination among residents might be an easy and cost-effective method to complement other promoting vaccination interventions. Also, this approach could be extended to other healthcare workers’ categories or even to general population. However, this study shows that a nontransparent nudge may simultaneously be effective and make nudged people feel they can control their choices. Therefore, we must be careful about the ethical implications when using defaults to nudge, especially since the sense of autonomy was found to be stronger among people on whom the nudge was effective.

## Supporting information

Annexes

## Data Availability

All data produced in the present study are available upon reasonable request to the authors

## Acknowledgements

We would like to thank the residents who participated, Pr. Michel Carles, Mme Aurore Schneider, Mr. Jerôme Chauvet, Dr. Philippe Colombani, Dr. Serge Tempesta and Dr. Astrid Sieber-Roth for allowing this study to be carried out, Pr. Michel Benoît, Pr. Michel Carles and Pr. Isabelle Milhabet for proofreading.

## Declaration

### Ethical approval

As an interventional study in the human and social sciences applied to the field of health, the study was classified out of scope of the Jardé law by the Ethics Committee “CPP Sud Ouest et Outre Mer III” (ID-RCB: 2021-A00975-36, Ref. SI CNRIPH: ID 12087 N°21.03.31.68613). A favorable opinion was issued by the Ethics Committee of the French National College of Generalist Teachers (N° 020921306).

## Data availability

The data underlying this article will be shared upon request to the corresponding author.

