## Supplementary material for "Effectiveness and acceptability of an opt-out nudge to promote influenza vaccination among medical residents in Nice, France: a randomized controlled trial": Annexes

**Annex 1**

*Telephone interview guide for the step 1 call*

"Good morning, I'm Ilaria Serati, a General Medicine resident. Are you Mr./Mrs. X?

As you surely know, the influenza vaccination campaign has begun. I may propose you a vaccination appointment the [day] at [hour] at [the vaccination center the resident depends on according to his/her internship site]. I would like to know if you accept this appointment, or if you want to reschedule or cancel it."

→ If the resident accepts: the appointment is scheduled and eventually transferred to the vaccination center, depending on the center working operation.

→ If the resident wants to reschedule the appointment: another appointment is offered.

→ If the resident has already been vaccinated or refuse the appointment: close the call cordially.

In any case, close the call cordially without mentioning the study.

**Annex 2**

*Telephone interview guide for the step 2 call*

"Good morning, I'm Ilaria Serati, a General Medicine resident. Are you Mr./Mrs. X?

I'm working on a thesis about influenza vaccination promotion, would you have five minutes to answer some questions?

By answering these questions, you agree that the data collected anonymously by this questionnaire will be subject to strictly confidential and secure computerizing processing for scientific purposes as part of a medical thesis. The data will be stored on an encrypted flash drive. Answering this questionnaire is not compulsory and you can leave the study at any time without having to justify your choice, the data collected concerning you will then be deleted. To do so, simply send a request by e-mail to the following address: or contact me by phone at [investigator's phone number]. The expected benefits of this research are to better understand how to motivate health professionals to vaccinate and what they consider acceptable. The project leader is Dr. Adriaan Barbaroux, general practitioner and teacher researcher at the Department of General Medicine."

→ Collection of verbal consent and completion of the questionnaire.

→ Close the call by thanking the participants for their participation and offer to send them the consent form and the results of the study by e-mail.

*Questionnaire*

Date of completion of the questionnaire:

Age, gender, year of graduation:

- Last year, did you get a flu shot?
- On January 9th, have you already got a flu shot?
- *[for the nudge group only]* If you were vaccinated, was it during an appointment I gave you last November?

- On a scale of 1 to 7 from 1 = "strongly disagree" to 7 = "strongly agree", how do you feel about the current vaccine recommendations?

Strongly disagree Strongly agree

1 2 3 4 5 6 7

- *[if not yet vaccinated only]* On a scale of 1 to 7 from 1 = "not at all" to 7 = "absolutely", do you intend to be vaccinated this year?

Not at all Absolutely

1 2 3 4 5 6 7

- Do you intend to be vaccinated in the following years?

Not at all Absolutely

1 2 3 4 5 6 7

- Would you advise other health care professionals to get vaccinated this year?

Not at all Absolutely

1 2 3 4 5 6 7

- Would you advise patients to get vaccinated this year?

Not at all Absolutely

1 2 3 4 5 6 7

- In a first step of this study, we scheduled some residents for a flu shot at the vaccination center. The residents could reschedule or cancel their appointment. This procedure was intended to push students to get vaccinated by giving them a nudge. Studies show that having an appointment made for you and offered up front increases the likelihood of getting vaccinated compared to having to make an appointment by yourself.

What do you think about the use of this type of method on you? On a scale of 1 to 7 from 1 = "not at all" to 7 = "absolutely", this type of procedure is:

| | Not at all | | | | | | Absolutely |
|---|---|---|---|---|---|---|---|
| Abusive | 1 | 2 | 3 | 4 | 5 | 6 | 7 |

| | | | | | | | |
|---|---|---|---|---|---|---|---|
| Acceptable | 1 | 2 | 3 | 4 | 5 | 6 | 7 |
| Inappropriate | 1 | 2 | 3 | 4 | 5 | 6 | 7 |
| Ethical | 1 | 2 | 3 | 4 | 5 | 6 | 7 |
| Immoral | 1 | 2 | 3 | 4 | 5 | 6 | 7 |
| Unfair | 1 | 2 | 3 | 4 | 5 | 6 | 7 |
| Legitimate | 1 | 2 | 3 | 4 | 5 | 6 | 7 |
| Relevant | 1 | 2 | 3 | 4 | 5 | 6 | 7 |

- What do you think about the use of this type of method on patients? On a scale of 1 to 7 from 1 = "not at all" to 7 = "absolutely", this type of procedure is:

| | Not at all | | | | | | Absolutely |
|---|---|---|---|---|---|---|---|
| Relevant | 1 | 2 | 3 | 4 | 5 | 6 | 7 |
| Legitimate | 1 | 2 | 3 | 4 | 5 | 6 | 7 |
| Unfair | 1 | 2 | 3 | 4 | 5 | 6 | 7 |
| Immoral | 1 | 2 | 3 | 4 | 5 | 6 | 7 |
| Ethical | 1 | 2 | 3 | 4 | 5 | 6 | 7 |
| Inappropriate | 1 | 2 | 3 | 4 | 5 | 6 | 7 |
| Acceptable | 1 | 2 | 3 | 4 | 5 | 6 | 7 |
| Abusive | 1 | 2 | 3 | 4 | 5 | 6 | 7 |

- Next year, would you like to be offered a vaccination appointment up front?

| Not at all | | | | | | Absolutely |
|---|---|---|---|---|---|---|
| 1 | 2 | 3 | 4 | 5 | 6 | 7 |

- Do you think this nudge lets you choose whether to get vaccinated?

| Not at all | | | | | | Absolutely |
|---|---|---|---|---|---|---|
| 1 | 2 | 3 | 4 | 5 | 6 | 7 |
